# The Set Point Is Not Where We Thought: The Primacy of Baroreflex Gain Variability

**DOI:** 10.64898/2026.03.23.26349128

**Authors:** Alexander Weaver, Aiden Yakimchuk, Ryan Woodman, Warren Lockette

## Abstract

For decades, cardiovascular physiology has been built on the assumption that arterial barorecep-tors adjust heart rate (HR) to maintain a defined blood pressure set point. We challenge this paradigm fundamentally. Blood pressure and heart rate *both* change substantially in response to physiological stress and neither returns reliably to a fixed baseline value. This raises the question of whether a higher-order variable, one that remains stable while blood pressure and heart rate reset freely might better represent a truly defended, “set-point” quantity. We hypothesized that instantaneous baroreceptor gain (IBS), expressed as the change in R-R interval per unit change in systolic blood pressure (SBP), has a coefficient of variation (IBS CV) that is invariant across different physiological challenges. If IBS CV is fixed, then ΔHR and ΔSBP must vary proportionally, maintaining a stable gain relationship even as each changes in magnitude. This constraint suggests that blood pressure and heart rate are adjustable outputs that reset freely in response to physiological demand, rather than defended targets in their own right. An invariant IBS CV guarantees that the baroreflex arc behaves consistently and predictably across the full range of physiological states: the brain can rely on a stable gain relationship between blood pressure and heart rate, enabling reliable closed-loop cardiovascular control without having to recalibrate the relationship under every new condition.

To test this hypothesis, we had healthy adult volunteers undergo either the cold pressor test or passive orthostatic challenge. Heart rate (HR), systolic blood pressure (SBP), IBS (the regression coefficient of R-R intervals on their respective systolic blood pressures), and the coefficients of variation (CV, i.e. standard deviation ÷ mean value) of each were measured at baseline and during each stress perturbation. During orthostatic challenge, HR rose significantly while SBP fell significantly. Classically, this HR rise is attributed to baroreflex compensation for falling pressure. However, the critical observation is that SBP was not restored to baseline. Instead, it remained substantially reduced while HR stayed persistently elevated and HR CV increased significantly. A system primarily defending a blood pressure set point should augment baroreflex gain and suppress pressure variability; instead mean IBS showed no significant change, SBP CV amplified more than threefold, and IBS CV was unchanged. During the cold pressor test, both HR and SBP rose simultaneously, which is inconsistent with a pressure-defending system that would have suppressed HR in response to the large rise in SBP. IBS CV was also stable across this perturbation while SBP CV amplified dramatically. These findings challenge the classical baroreceptor set-point model and suggest that IBS CV, not blood pressure, is the primary regulated cardiovascular variable.

## Introduction

The most deeply entrenched assumption in cardiovascular physiology that has shaped textbook teaching, clinical practice, and decades of research is that arterial baroreceptors define a blood pressure set point, and that heart rate is a secondarily adjusted variable that remediates deviations from that blood pressure set point.^**1,2**^ Under this classical negative-feedback model, a rise in blood pressure activates carotid sinus and aortic arch baroreceptors which signal the nucleus tractus solitarius to reduce efferent sympathetic tone and increase parasympathetic outflow and consequently slows the heart until pressure returns toward the defended level.^**3,4**^ This model has been extraordinarily influential, yet its explanatory scope is narrower than is commonly appreciated. It only accommodates conditions in which HR and blood pressure move in opposite directions, but it fails to explain conditions in which these cardiovascular variables move in the same direction, or more commonly, conditions in which the predicted pressure restoration simply does not occur despite a large HR response.

During orthostatic stress, blood pressure falls and heart rate rises. This response represents the classical model of baroreflex compensation. Yet, the blood pressure is never restored to its baseline and the baroreflex gain (IBS) does not increase to defend it which exposes the limits of a pressure-as-set-point account.^**5,6**^ During vigorous exercise or cold pressor pain both heart rate and blood pressure rise together.^**7**^ These observations are not anomalies to be explained away; they are the rule. The baroreceptor-as-set-point model accommodates them only by invoking ad hoc mechanisms such as baroreflex resetting, central command override, and efference copy that progressively hollow out the explanatory core of the original theory.

We propose a fundamentally different framework. We argue that the coefficient of variation of instantaneous baroreceptor gain (IBS CV) is the true cardiovascular set point. IBS CV is the ratio of the standard deviation of IBS to its mean; its invariance means that this ratio is a stable, individual-specific property that is unchanged across physiological challenge. Mean heart rate, blood pressure, or IBS itself may shift under different conditions, but the ratio of IBS’s standard deviation to its mean does not. The functional importance of this invariance cannot be overstated: a fixed IBS CV ensures that the baroreflex arc maintains a consistent and predictable gain relationship between blood pressure and heart rate across all physiological states. Blood pressure and heart rate are therefore free to reset to new values, while IBS CV remains the defended set point. Blood pressure, in this framework, is a consequential downstream output and not the primary variable being defended.^**8,9**^

This reframing has been hindered historically for many reasons. The methods required to test this hypothesis—continuous beat-to-beat blood pressure monitoring, computational sequence analysis to extract instantaneous baroreflex sensitivity and statistical assessment of gain variability across physiological states were unavailable until recent decades. Even with these tools now accessible, the methodology remains more complex than traditional baroreflex assessment. Rather than focusing on first-order variables such as mean blood pressure or heart rate, it requires analysis of second-order statistics, specifically the variability (coefficient of variation) of a dynamic gain parameter over time, demanding both technical sophistication and a conceptual shift away from classical set-point thinking. The dominance of blood pressure as the assumed regulated variable also arose from practical considerations: pressure is far easier to measure non-invasively than stroke volume or ventricular filling pressure, which biased the experimental literature toward pressure-centric frameworks.^**10**^ The anatomical discreteness of the aortic and carotid baroreceptor pathway also formed a clean, describable arc from stretch receptor to brainstem nucleus to effector organ and made it an irresistible target for classical physiological analysis.^**11,12**^

Using two physiological perturbations, the cold pressor stress and orthostatic challenge, we expose the relationship between HR, SBP, IBS, and their respective variability in conditions that differentially engage the sympathetic nervous system and Frank–Starling mechanics.

## Methods

The study was approved by our institutional committee for the use of human subjects in research, and all subjects gave written informed consent. Two cohorts of healthy adult volunteers participated under resting baseline conditions followed by a physiological challenge. The cold pressor cohort comprised 14 subjects with complete HR and SBP data (13 with complete IBS data). The orthostatic cohort comprised 16 subjects with complete HR and SBP data (9 with complete IBS data). All subjects rested supine for a minimum of ten minutes before baseline measurements. For cold pressor testing, subjects immersed the non-dominant forearm in ice water (0°C) for up to three minutes. For orthostatic challenge, subjects transitioned from supine to unsupported standing, with measurements continuing for at least three minutes upright. Heart rate (R-R intervals) and SBP were continually measured beat-by-beat using non-invasive finger photoplethysmometry (Finapres). IBS was computed as the linear regression coefficient (ms/mmHg) of the R-R interval on SBP over sequential three-minute epochs using the sequence method, a validated technique that identifies consecutive beats in which SBP and R-R interval change in the same direction and calculates their regression slope.^**13**^ The CV for each variable was computed within each epoch as (standard deviation ÷ mean) × 100%. Within-subject differences between baseline and challenge conditions were assessed by paired two-tailed t-tests. Relationships between R-R interval and IBS across subjects and conditions were assessed by Pearson correlation with corresponding t-statistics. Data are expressed as mean ± SD. Statistical significance was set at α = 0.05. Sample sizes were determined prospectively based on the primary outcome of interest: the within-subject change in IBS CV across physiological challenge. From published values of instantaneous baroreflex sensitivity in healthy adults, the coefficient of variation of IBS is approximately 40–100%, with within-subject test-retest standard deviations of roughly 25–35 percentage points. To detect a meaningful change in IBS CV of 20 percentage points — a conservative threshold representing one-half standard deviation of the baseline distribution — with 80% power at a two-tailed α of 0.05 using a paired t-test, a minimum of 12 subjects is required. Both the cold pressor cohort and the orthostatic cohort (n = 16 for HR and SBP) met or exceeded this threshold for their primary comparisons.

## Results

During the cold pressor test, heart rate increased significantly from rest (68.2 ± 11.1 bpm) to cold immersion (73.8 ± 12.1 bpm; t(13) = −3.46, p < 0.01; Figure 1A). Systolic blood pressure rose dramatically from rest (115.3 ± 20.3 mmHg) to cold pressor (144.9 ± 18.9 mmHg; t(13) = −8.14, p < 0.001; Figure 1B). Critically, both HR and SBP rose simultaneously. If baroreceptors were defending a blood pressure set point, the reflex response to rising SBP should have suppressed HR. Instead, HR rose. This simultaneous co-elevation of HR and SBP under cold stress is a direct empirical challenge to the baroreceptor setpoint model and is precisely what is predicted if the primary regulated variable is cardiac output (maintained here by both tachycardia and increased vascular resistance) rather than arterial pressure per se.

**Figure 1.**
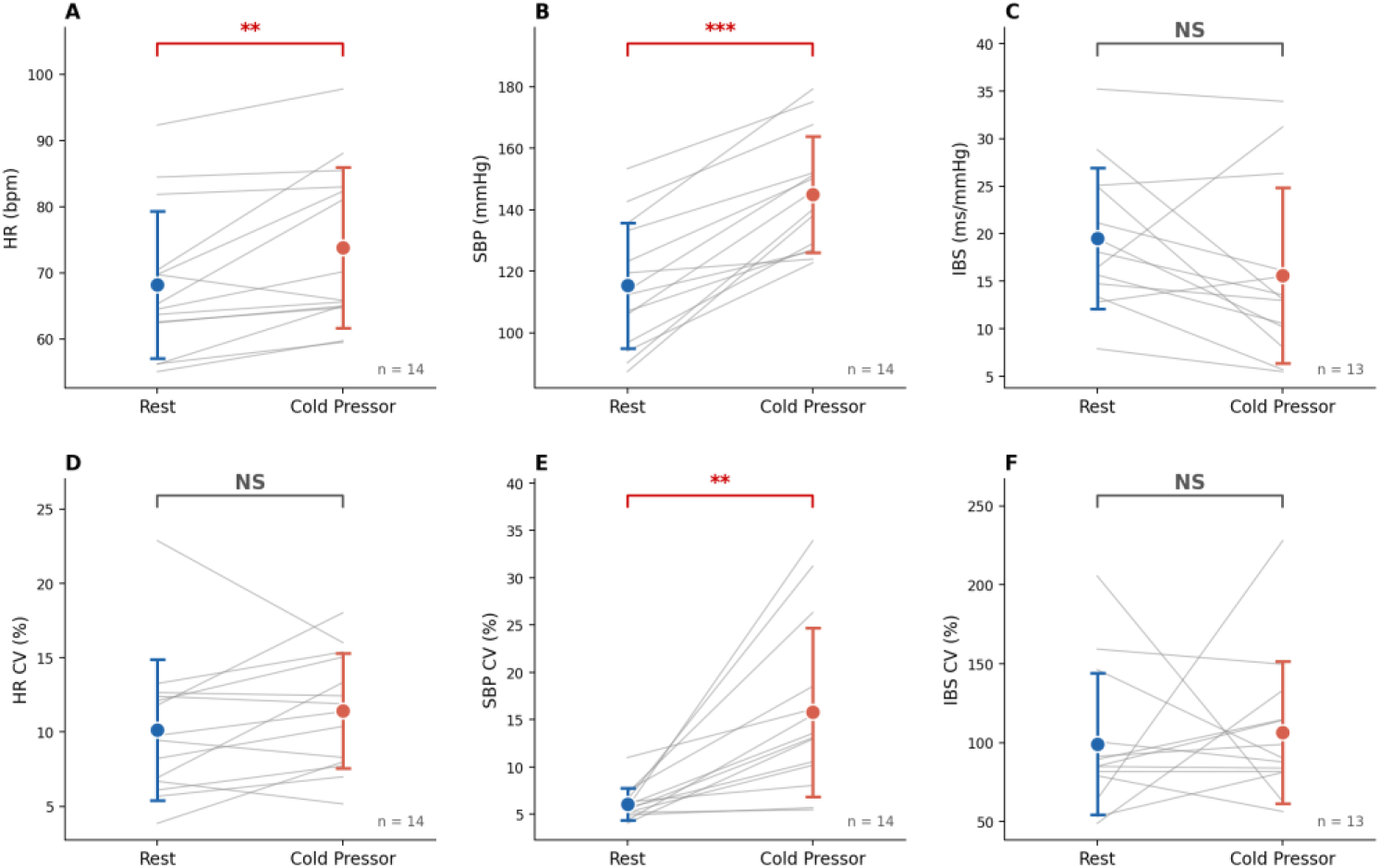
Cold Pressor Test: Paired measurements of HR (A), SBP (B), IBS (C), HR CV (D), SBP CV (E), and IBS CV (F) at rest and during cold pressor. Filled circles = group mean ± SD. Gray lines = individual paired responses. *, **, *** indicate p < 0.05, 0.01, 0.001 respectively; NS = not significant.

There were no significant differences in IBS between resting and cold pressor (19.5 ± 7.4 to 15.6 ± 9.2 ms/mmHg; t(12) = 1.74, NS; Figure 1C), suggesting that baroreflex gain was not augmented to buffer the large SBP rise, the opposite of what a pressure-defending system would be expected to do. The CV of HR was preserved (10.1 ± 4.7% to 11.4 ± 3.9%; NS; Figure 1D) while the CV of SBP more than doubled (6.1 ± 1.7% to 15.8 ± 8.9%; p < 0.01; Figure 1E).

Importantly, the CV of IBS was similarly unchanged (99.2 ± 45.0% vs. 106.3 ± 45.0%; NS; Figure 1F). The pattern of preserved HR variability, dramatically amplified SBP variability, and stable baroreflex gain variability is inconsistent with pressure being the primary defended variable and consistent with the system defending the variability structure of the baroreceptor gain. This pattern of dramatically amplified SBP variability alongside an unchanged IBS CV is precisely what is expected if IBS CV is the defended set point.

During the orthostatic challenge, HR rose substantially (64.3 ± 12.8 to 81.7 ± 11.8 bpm; t(15) = −10.00, p < 0.001; Figure 2A) while SBP fell significantly (115.2 ± 10.3 to 95.9 ± 19.4 mmHg; t(14) = 4.78, p < 0.001; Figure 2B). IBS declined numerically from supine to upright (9.5 ± 3.6 to 6.7 ± 4.1 ms/mmHg; t(8) = 1.68, NS; Figure 2C). Classically, this HR rise is interpreted as baroreflex compensation for falling pressure. A system primarily defending a blood pressure set point should also increase baroreflex gain, restore pressure, and tighten pressure variability. None of these occur.^**5,6**^ The CV of HR increased significantly from supine to upright (5.8 ± 3.1% to 8.6 ± 3.3%; t(15) = −2.40, p < 0.05; Figure 2D), indicating that the autonomic system is exploring a broader range of cardiac states and sampling more widely from the HR state space as it locates the new Frank–Starling operating point.^**8,9,14**^ The CV of SBP increased dramatically (4.7 ± 1.4% to 15.9 ± 9.0%; t(15) = −4.83, p < 0.001; Figure 2E), a more than threefold amplification, while IBS CV was again stable (68.3 ± 24.1% vs. 70.4 ± 34.0%; NS; Figure 2F). The pattern of HR rising as SBP falls with HR and SBP variability increasing while IBS variability is stable fits a coherent picture in which IBS CV is the defended quantity, and stroke volume accommodates the physiological demand via Frank–Starling mechanics, and blood pressure finds its own equilibrium as a downstream consequence.

**Figure 2.**
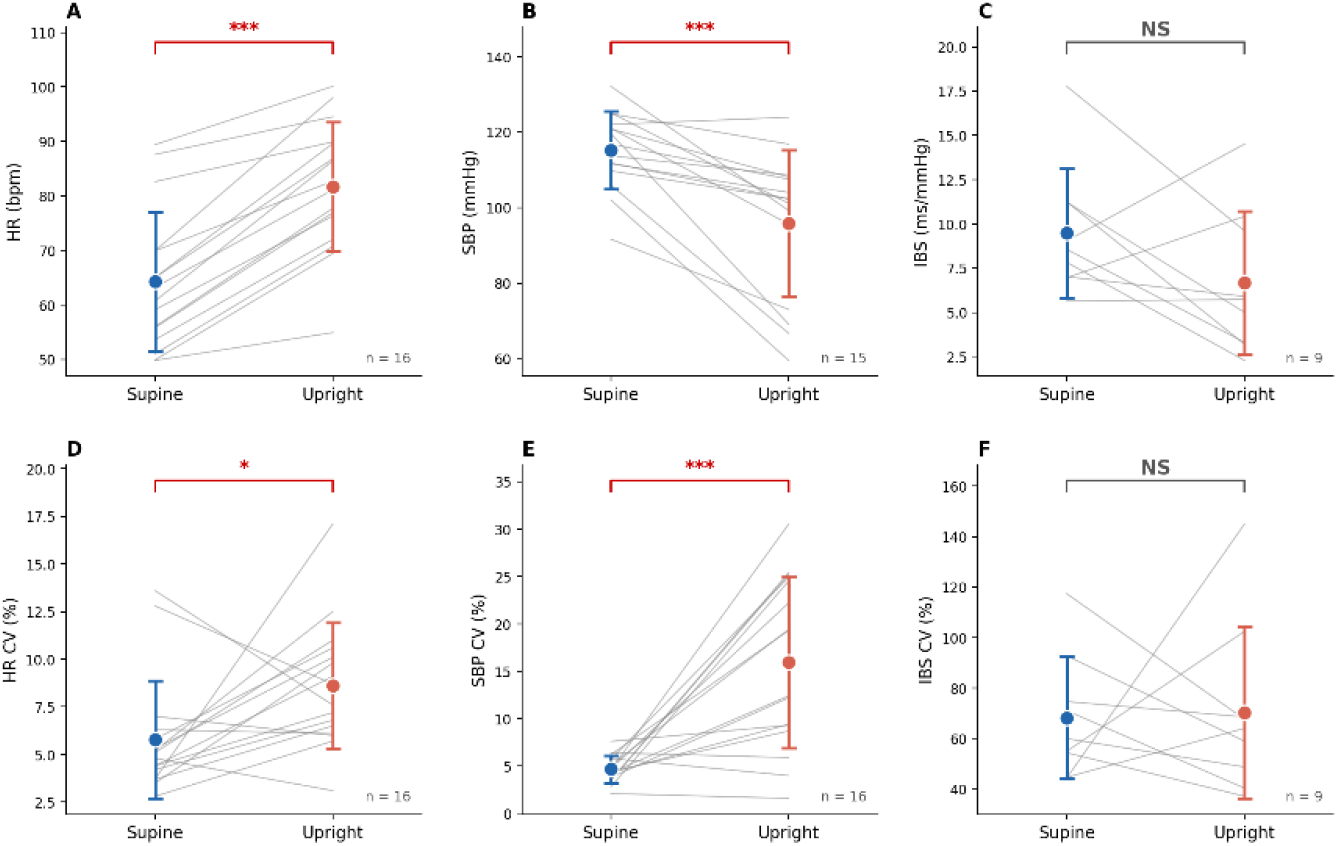
Orthostatic Challenge: Paired measurements of HR (A), SBP (B), IBS (C), HR CV (D), SBP CV (E), and IBS CV (F) while supine and upright. Filled circles = group mean ± SD. Gray lines = individual paired responses. Significance notation as in Figure 1.

Figure 3 plots R-R interval against IBS on a logarithmic y-axis for both stressor conditions. As R-R interval shortens, IBS falls under both cold pressor (n = 14; r = 0.679, p = 0.008) and orthostatic challenge (n = 30; r = 0.626, p < 0.001). The slopes of the two regression lines are statistically indistinguishable (slope difference p = 0.80), indicating that the relationship between cardiac interval and IBS is preserved across two mechanistically distinct stressors in which SBP moved in opposite directions. The cold pressor relationship is shifted upward, with higher IBS at any given R-R interval. Despite opposite SBP responses across the two stressors, the structural relationship between cardiac interval and baroreflex gain is preserved.

**Figure 3.**
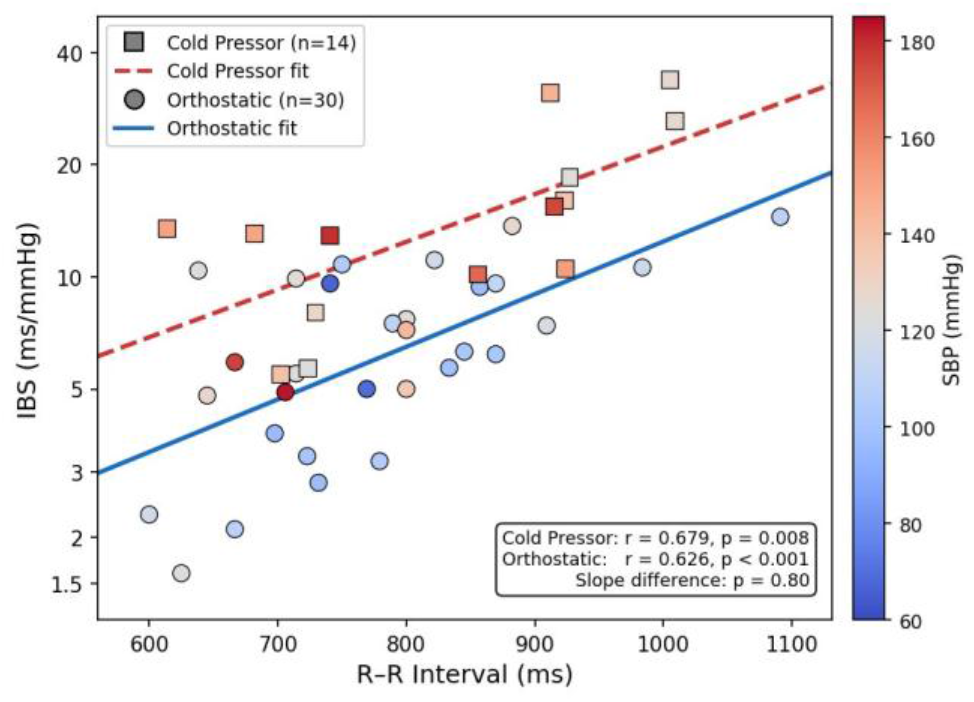
R-R interval plotted against instantaneous baroreflex sensitivity (IBS) on a logarithmic y-axis for cold pressor (squares, dashed regression line, n = 14) and orthostatic challenge (circles, solid regression line, n = 30). The color of each point indicates the systolic blood pressure (SBP) recorded at that observation, ranging from deep blue (lowest SBP, approximately 60 mmHg) through white (approximately 120 mmHg) to deep red (highest SBP, approximately 180 mmHg), as shown in the color bar at right. The two stressors produce opposite SBP responses: cold pressor raises both heart rate and SBP, producing predominantly warm-colored squares; orthostasis raises heart rate but lowers SBP, producing predominantly cool-colored circles. Despite this divergence in SBP direction, IBS tracks R-R interval similarly under both conditions. As R-R interval shortens (heart rate rises), IBS falls in both stressor conditions, and the slopes of the two regression lines are statistically indistinguishable (Cold Pressor: *r* = 0.679, *p* = 0.008; Orthostatic: *r* = 0.626, *p* < 0.001; slope difference *p* = 0.80). The cold pressor line sits above the orthostatic line at every R-R interval, reflecting higher IBS under cold stress independent of heart rate. The identical slopes across two conditions in which SBP moved in opposite directions demonstrate that the structural relationship between heart rate and IBS is preserved regardless of SBP level or direction; SBP varies freely while this relationship does not. The IBS-to-heart-rate relationship is a stable physiological structure across mechanistically distinct stressors.

Figure 4 examines the scaling relationship between the standard deviation and mean of IBS across all subject-condition data points (n = 40) from both cohorts. On logarithmic axes, the fitted slope of SD on mean IBS was 1.140 (95% CI: 0.936–1.344, r^2^ = 0.771, p < 0.001), and a formal test did not reject a slope of 1 (p = 0.174). When the standard deviation of a variable scales proportionally with its mean, the coefficient of variation is constant. These data indicate that IBS CV is approximately constant across subjects, conditions, and stressor types.

**Figure 4.**
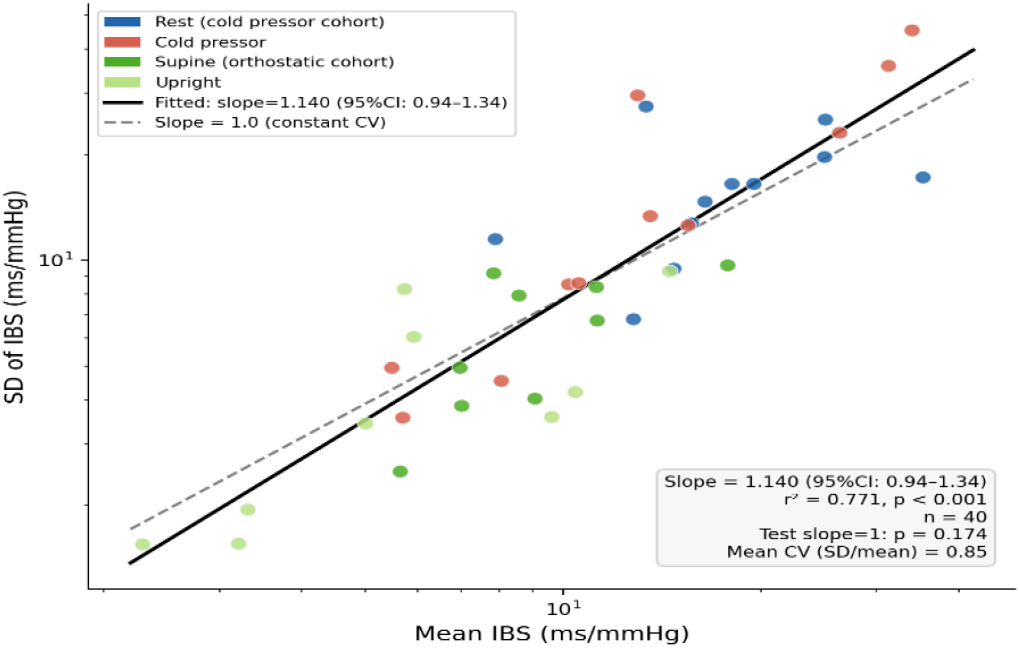
Scaling relationship for instantaneous baroreflex sensitivity. The standard deviation (SD) of IBS is plotted against mean IBS on logarithmic axes for each subject-condition data point (n = 40) from the cold pressor cohort (rest and cold pressor, blue and red circles) and orthostatic cohort (supine and upright, dark and light green circles). When SD is proportional to mean, i.e., when the CV is constant, the slope of this log-log plot equals 1. The fitted slope was 1.140 (95% CI: 0.936–1.344, r^2^ = 0.771, p < 0.001), and a formal test cannot reject a slope of 1 (p = 0.174). The dashed line shows the theoretical prediction of slope = 1. SD scales proportionally with mean, and the CV is approximately constant across all conditions tested. Because IBS CV is the defended set point, SD scales proportionally with mean as a necessary consequence, and this structure is preserved across all conditions in which the intact baroreflex arc is functioning.

## Discussion

The assumption that baroreceptors define the cardiovascular set point has been so thoroughly integrated into physiological teaching that it has acquired the status of established fact rather than theoretical assumption.^**1,2,3**^ Yet, as inferred from the present data, the baroreceptor set-point model fails important empirical tests when examined carefully. The established model predicts that when blood pressure falls, heart rate should rise to restore it, and conversely, when blood pressure rises, heart rate should fall. The orthostatic data superficially satisfy the first prediction: SBP fell and HR rose. But the model’s deeper prediction—that this reflex should restore blood pressure toward its baseline—is not fulfilled. SBP remained substantially reduced throughout the upright period, mean IBS showed no significant change to augment the pressure-recovery reflex, and SBP CV amplified threefold rather than being suppressed. The cold pressor data provide an even more direct challenge: both HR and SBP rose together by 5.6 bpm and approximately 30 mmHg. A system defending a blood pressure set point should have suppressed HR as SBP rose sharply. Instead, HR accelerated. This co-elevation of HR and SBP is incompatible with baroreceptor-mediated pressure defense and is the clearest single observation in our dataset against the classical model.

What the data support is a system in which IBS CV is the defended quantity, with heart rate and blood pressure free to reset while stroke volume absorbs physiological demand through Frank–Starling mechanics. This is not merely a semantic reframing. If IBS CV is the defended variable, then (1) IBS CV should remain stable across physiological perturbations while mean IBS, HR, and SBP shift freely; (2) SBP variability should increase, not decrease, during stress because pressure is a free output rather than a tightly regulated target; and (3) the hierarchical stability of CVs should follow regulatory priority, with the defended variable (IBS CV) showing maximal resistance to perturbation. All three predictions are confirmed in our data. *The most parsimonious interpretation is that what remains invariant under perturbation is the defended variable*.

Defending IBS CV while allowing heart rate and blood pressure to reset independently requires a mechanical degree of freedom. The Frank–Starling mechanism facilitates this defense by providing the necessary volumetric flexibility. If IBS CV is fixed, and if stroke volume were also fixed, then HR and SBP would be mechanistically locked together. By allowing stroke volume to vary freely with filling pressure, the Frank–Starling relationship provides the additional degree of freedom necessary for IBS CV to remain invariant while HR and SBP reset independently. Stroke volume adjusts automatically in response to venous return so that when physiological demand increases during orthostasis or cold stress, heart rate and blood pressure can adjust to whatever values maintain the stable IBS CV ratio. Without this volumetric freedom, a fixed stroke volume would force HR and SBP to vary in strict lockstep, making independent adjustment of each impossible and precluding the dissociation between their respective CVs that our data demonstrate: it is precisely because stroke volume is free to vary that IBS CV can remain invariant while HR and SBP reset to entirely different values under each stressor.The plausibility of this framework requires that the central nervous system can track the beat-by-beat variability of baroreflex gain. To accomplish this, the necessary neural architecture exists at every level of the cardiovascular afferent pathway, from arterial baroreceptor firing through brainstem integration to cortical representation.

Arterial baroreceptors transduce each pressure pulse into action potential trains whose inter-spike intervals encode both the magnitude and rate of rise of the pressure wave, so that the signal arriving at the nucleus tractus solitarius (NTS) carries information about the variability of the pressure waveform, not just its mean.^**15**^ At the first NTS synapse, frequency-dependent synaptic depression causes the postsynaptic response to be largest when the input pattern changes and smallest when it is sustained, effectively making this synapse a detector of shifts in baroreceptor firing rather than a reporter of average firing rate.^**16,17**^ Critically, most second-order NTS neurons receive convergent input from both arterial baroreceptors and cardiac mechanoreceptors, positioning them to encode the beat-by-beat relationship between R-R interval and systolic blood pressure on every cardiac cycle which is precisely the instantaneous baroreflex slope.^**18,19**^

Cardiac sympathetic afferents provide an independent and parallel channel of variability information.^**20**^ These afferents encode ventricular wall tension and filling volume rather than arterial pressure, and their firing patterns carry information about beat-by-beat stroke volume variability through the Frank– Starling relationship. Their convergence with arterial baroreceptor afferents at the NTS gives this nucleus the two raw inputs — pressure timing and cardiac filling that are needed to represent the variability of the baroreflex gain relationship. The parabrachial nucleus serves as a critical relay in this pathway, receiving cardiac mechanoreceptor input encoding myocardial stretch and projecting it to higher cortical centers.^**21,22**^ In functional terms, it transmits a volume and filling error signal that represents the Frank–Starling state of the heart directly, completing the afferent information the cortex requires to track IBS variability.

The insular cortex is the primary cortical node for the overall integration of this function.^**23**^ The posterior insula receives both cardiac timing signals encoding the R-R interval and blood pressure afferents relayed from the NTS through the parabrachial nucleus and the ventromedial posterior thalamic nucleus.^**24**^ Because these two signals arrive together with each cardiac cycle, the insular cortex is positioned to compute their moment-to-moment regression relationship (i.e., the IBS) and to track the variability of that gain over time.^**25**^ The anterior insula translates this representation into a prediction about what the IBS CV should be, given the individual’s autonomic architecture and top-down contextual priors.^**21,22,26**^ Deviations from this prediction, a shift in IBS CV away from the individual’s characteristic value, generate prediction errors that propagate to the anterior cingulate cortex and prefrontal cortex, updating the generative model and, through descending projections via the hypothalamus and brainstem, adjusting efferent autonomic commands.

Predictive coding is a computational framework in which the brain maintains an internal model of expected sensory input and continuously compares that prediction against what it actually receives; the confidence the brain assigns to any mismatch is called precision.^**27,28,29**^ In this framework, IBS CV maps directly onto the precision of baroreceptor prediction errors, the weight the brain assigns to beat-by-beat mismatches between expected and actual cardiovascular input. A stable IBS CV means that this weighting does not change under physiological challenge. When IBS CV matches the brain’s expected value, prediction errors remain small and no corrective efferent command is issued. When IBS CV deviates, prediction errors grow, and descending autonomic commands adjust to restore it.^**14**^ The brain does not compute IBS CV as an explicit number. Rather, it is encoded implicitly as the fixed confidence weighting in the brain’s cardiovascular model, and its defense is the natural consequence of a system that minimizes prediction errors scaled by that weighting. This fixedness is the computational signature of a set point: heart rate and blood pressure are free to vary because they are the outputs the system adjusts to keep IBS CV constant, not the quantities being defended.

The three CVs in our dataset behave in a clearly hierarchical fashion under physiological challenge, and this hierarchy is itself a central finding. SBP CV is maximally sensitive to perturbation, HR CV is intermediately sensitive, and IBS CV is completely insensitive—unchanged in both experiments, consistent with our observation and that of others^**30**^ that the coefficient of variation of IBS is invariant across many maneuvers. The correct interpretation of this hierarchy is one of *regulatory priority*. The variable that changes most freely is under least regulatory constraint: blood pressure. The variable that changes moderately is under intermediate constraint: heart rate. The variable that does not change at all is under maximal constraint: IBS CV. A system designed to defend a set point should show maximal resistance to perturbation in the defended variable, and that is precisely what IBS CV exhibits. The fact that SBP CV amplifies so dramatically is not evidence that blood pressure is poorly regulated; it is evidence that blood pressure is *free* to find whatever value is needed to maintain the invariant IBS CV.

Our finding that IBS CV is invariant across stressors is consistent with the independent work of others^**30**^ who demonstrated that the CV of IBS is invariant across a wide range of physiological maneuvers. Those findings, combined with our own data from two mechanistically distinct stressors, strongly suggest that IBS CV is not a dynamic state variable that the cardiovascular system adjusts in response to challenge, but a fixed individual characteristic, a constitutional property of each person’s autonomic architecture that is defended under perturbation in the manner expected of a true set point.

The individual-level data from Figure 4 clarify the interpretation of this scaling. Individual mean IBS shifts substantially and inconsistently across stressors, with no reliable preservation of rank order between conditions, whereas IBS CV does not change. This dissociation is decisive: mean IBS is not stable within individuals and is therefore not the defended quantity; IBS CV is stable within individuals and therefore is. Others have reported a constant SD/mean ratio of approximately 0.40 using 10-second sliding windows; we find a mean CV of approximately 0.85 using 3-minute epochs. This difference is expected: longer epochs average out more moment-to-moment variation and yield a higher apparent CV. Despite this difference in absolute level, both datasets show the same structural property: when the CV is constant, the standard deviation must rise and fall in direct proportion to the mean.

The biological authenticity of this scaling relationship, rather than it being a mathematical artifact of the CV calculation or a consequence of limited sample size, was confirmed by testing a null model in which fifteen sets of 12 random integers drawn from a uniform distribution (range 1–100) were subjected to identical log-log regression analysis. The null model yielded a non-significant slope (r = −0.439, p = 0.100), confirming that the analytical framework does not generate spurious Taylor’s Law scaling from data that contain no biological structure. The significant slope observed in the IBS data therefore reflects genuine physiological constraint, not mathematical artifact. This places IBS CV among a class of conserved biological scaling quantities that are mathematical in expression but biological in origin analogous to the Fibonacci sequence, which is not imposed on living systems by mathematics but emerges from them because a universal optimization constraint (minimization of energy, space, and structural redundancy) finds its natural expression in a recursive numerical relationship. Taylor’s Power Law appears across ecology, neuroscience, genomics, and cardiovascular physiology for the same reason: it is the mathematical fingerprint of a system operating under a defended, hierarchical constraint on variability, not an artifact of the analysis used to reveal it.^**31,32**^ This scaling likely represents an active neural defense of a proportional gain relationship. Data from denervation of the baroreflex in rodents suggest active defense rather than passive constraint. Sinoaortic denervation studies in conscious rats demonstrate that complete removal of baroreceptor afferents renders IBS unmeasurable while dramatically increasing the variances of both BP and IBI despite unchanged means,^**33**^ a fundamental disruption of the scaling architecture present in the intact baroreflex system. If IBS CV stability were merely a passive mathematical consequence of baroreflex dynamics, it should persist after denervation. The fact that it does not suggests the scaling requires intact neural circuitry and is therefore likely to be actively maintained. The proportional relationship of IBS SD to mean requires intact afferent innervation and is not preserved when the baroreflex arc is severed. The most direct test would be serial measurement of IBS mean and SD in cardiac transplant recipients as reinnervation progresses, in patients with graded autonomic neuropathy, and under pharmacological autonomic blockade.

Several limitations of the present study should be acknowledged. First, sample sizes are modest. However, the adequacy of sample size for the primary null finding of stability of IBS CV was confirmed post hoc: the cold pressor cohort yielded a within-subject mean difference in IBS CV of 7.1 percentage points with a standard deviation of approximately 45 percentage points, producing an observed effect size of Cohen’s d = 0.16 and achieved power of approximately 0.12, confirming that IBS CV was genuinely stable rather than undetectable due to insufficient power for that comparison. In contrast, the comparisons that were expected to reach significance, i.e., changes in HR, SBP, and SBP CV, achieved effect sizes of Cohen’s d = 0.47 to 1.95 across both the cold pressor and orthostatic cohorts, yielding power of 0.72 to >0.99, confirming that the null results for IBS CV were not attributable to inadequate statistical sensitivity in the overall study design. For the Pearson correlation analysis in Figure 4, a pooled sample of n = 40 subject-condition data points from both cohorts provides 80% power to detect a correlation of r ≥ 0.39 at α = 0.05; the observed r^2^ = 0.771 substantially exceeds this threshold. These data are appropriate for a preliminary study establishing the existence and direction of the described physiological relationships, and the present findings are intended to inform sample size requirements for a confirmatory investigation. Replication in larger, more diverse cohorts will be essential before the present findings can be considered definitive. Second, our participants are a convenience sample of healthy adults, and generalizability to clinical populations remains to be established. Third, the theoretical framework of predictive coding and active inference, while providing a coherent organizational structure, has not been directly tested here; the present findings are consistent with that framework but do not uniquely confirm it.

There are tremendous clinical implications for our reported findings: (1) A patient whose pressure is pharmacologically controlled but whose IBS CV has been disrupted may remain at elevated cardiovascular risk in ways that current monitoring is insufficient. (2) IBS CV offers a potentially more specific window into baroreflex arc integrity than either heart rate variability or baroreflex sensitivity alone because it captures the stability of the gain relationship rather than its magnitude. Serial IBS CV measurement in heart failure patients may prove a more sensitive marker of autonomic deterioration and a more meaningful therapeutic target than variables currently monitored. (3) The clinical significance of reduced HRV may depend critically on whether IBS CV is maintained. A patient with reduced HRV but preserved IBS CV occupies a fundamentally different physiological position than one in whom both are disrupted, and incorporating IBS CV into risk stratification may improve its specificity. (4) IBS CV disruption may represent a more sensitive and mechanistically specific marker of early baroreflex arc failure than heart rate or blood pressure variability alone, remaining informative even when mean baroreflex sensitivity has declined to the point where traditional measurements are unreliable.

## Data Availability

Anonymized data will be provided to investigators if requested in writing and if the use is intended for noncommercial purposes.

## References

1. Cowley AW Jr. Long-term control of arterial blood pressure. Physiol Rev. 1992;72(1):231–300.

2. Guyton AC, Coleman TG, Granger HJ. Circulation: overall regulation. Annu Rev Physiol. 1972;34:13–46.

3. Chapleau MW, Hajduczok G, Abboud FM. Mechanisms of resetting of arterial baroreceptors: an overview. Am J Med Sci. 1988;295(4):327–334.

4. Eckberg DL, Sleight P. Human Baroreflexes in Health and Disease. Oxford: Clarendon Press; 1992.

5. Wieling W, Krediet CT, van Dijk N, Linzer M, Tschakovsky ME. Initial orthostatic hypotension: review of a forgotten condition. Clin Sci. 2007;112(3):157–165.

6. Stewart JM. Mechanisms of sympathetic regulation in orthostatic intolerance. J Appl Physiol. 2012;113(10):1659–1668.

7. Rowell LB. Human Cardiovascular Control. New York: Oxford University Press; 1993.

8. Starling EH. The Linacre Lecture on the Law of the Heart. London: Longmans Green; 1918.

9. Katz AM. Ernest Henry Starling, his predecessors, and the Law of the Heart. Circulation. 2002; 106(23): 2986–2992.

10. Pickering TG, Hall JE, Appel LJ, et al. Recommendations for blood pressure measurement in humans and experimental animals. Circulation. 2005;111(5):697–716.

11. Andresen MC, Kunze DL. Nucleus tractus solitarius: gateway to neural circulatory control. Annu Rev Physiol. 1994;56:93–116.

12. Summers J, Miklja Z, Weaver A, Taha J, Yakimchuk A, Kramm C, Li Y, Taha W, Afonso L, Farrow S, Woodman R, Lockette W. Cardiac PIEZO 1 channels modulate anxiety. doi: 10.64898/2026.02.18.706706

13. Parati G, Di Rienzo M, Bertinieri G, Pomidossi G, Casadei R, Groppelli A, Pedotti A, Zanchetti A, Mancia G. Evaluation of the baroreceptor-heart rate reflex by 24-hour intra-arterial blood pressure monitoring in humans. Hypertension. 1988 Aug;12(2):214–22.

14. Pezzulo G, Rigoli F, Friston K. Active inference, homeostatic regulation and adaptive behavioural control. Prog Neurobiol. 2015;134:17–35.

15. Seagard JL, van Brederode JF, Dean C, Hopp FA, Gallenberg LA, Kampine JP. Firing characteristics of single-fiber carotid sinus baroreceptors. Circ Res. 1990;66(6):1499–1509.

16. Kline DD. Plasticity in glutamatergic NTS neurotransmission. Respir Physiol Neurobiol. 2008;164(1-2):105–111. doi:10.1016/j.resp.2008.04.

17. Andresen MC, Yang M. Dynamics of sensory afferent synaptic transmission in aortic baroreceptor regions of nucleus tractus solitarius. J Neurophysiol. 1995;74(4):1518–1528.

18. Simms AE, Paton JF, Pickering AE. Disinhibition of the cardiac limb of the arterial baroreflex in rat: a role for metabotropic glutamate receptors in the nucleus tractus solitarii. J Physiol. 2006 Sep 15;575(Pt 3):727–38. doi: 10.1113/jphysiol.2006.112672.

19. Seagard JL, Dean C, Hopp FA. Role of glutamate receptors in transmission of vagal cardiac input to neurones in the nucleus tractus solitarii in dogs. J Physiol. 1999;520(1):243–253.

20. Armour JA. Cardiac neuronal hierarchy in health and disease. Am J Physiol Regul Integr Comp Physiol. 2004;287(2):R262–271.

21. Saper CB, Loewy AD. Efferent connections of the parabrachial nucleus in the rat. Brain Res. 1980;197(2):291–317.

22. Palmiter RD. The parabrachial nucleus: CGRP neurons function as a general alarm. Trends Neurosci. 2018;41(5):280–293.

23. Craig AD. How do you feel — now? The anterior insula and human awareness. Nat Rev Neurosci. 2009;10(1):59–70.

24. Saper CB. The central autonomic nervous system: conscious visceral perception and autonomic pattern generation. Annu Rev Neurosci. 2002;25:433–469.

25. Critchley HD, Wiens S, Rotshtein P, Ohman A, Dolan RJ. Neural systems supporting interoceptive awareness. Nat Neurosci. 2004;7(2):189–195.

26. Barrett LF, Simmons WK. Interoceptive predictions in the brain. Nat Rev Neurosci. 2015;16(7):419–429.

27. Friston K. The free-energy principle: a unified brain theory? Nat Rev Neurosci. 2010;11(2):127–138.

28. Clark A. Whatever next? Predictive brains, situated agents, and the future of cognitive science. Behav Brain Sci. 2013;36(3):181–204.

29. Seth AK, Friston KJ. Active interoceptive inference and the emotional brain. Philos Trans R Soc Lond B Biol Sci. 2016;371(1708):20160007.

30. Wesseling KH, Karemaker JM, Castiglioni P, et al. Validity and variability of xBRS: instantaneous cardiac baroreflex sensitivity. Physiol Rep. 2017;5(22):e13509. doi:10.14814/phy2.13509

31. Taylor LR. Aggregation, variance and the mean. Nature. 1961;189:732–735.

32. Livio M. The Golden Ratio: The Story of Phi, the World’s Most Astonishing Number. New York: Broadway Books; 2002.

33. Radaelli A, Mancia G, De Carlini C, Soriano F, Castiglioni P. Patterns of cardiovascular variability after long-term sino-aortic denervation in unanesthetized adult rats. Sci Rep. 2019;9(1):1232.

